# Isosorbide mononitrate / cilostazol for lacunar cerebral small vessel disease-outcomes at 6 months in the LACI-2 trial

**DOI:** 10.1101/2025.08.04.25332996

**Authors:** Philip M Bath, Lisa J Woodhouse, Iris Mhlanga, John Bamford, Vera Cvoro, Fergus N Doubal, Timothy J England, Ahamad Hassan, Alan A Montgomery, John T O’Brien, Christine Roffe, Nikola Sprigg, David J Werring, Joanna M Wardlaw, LACI-2 Trial Investigators

**Author notes:** LACI-2 Investigators are listed in Appendix. Correspondence to: Prof Philip Bath Stroke Trials Unit, University of Nottingham Queen’s Medical Centre D Floor, South Block Derby Road, Nottingham NG7 2UH UK.

## Abstract

**Background:** Lacunar stroke, a type of cerebral small vessel disease (cSVD), causes cognitive decline and dependency. The LACunar Intervention Trial-2 (LACI-2) trial showed that 12 months of treatment with isosorbide-mononitrate (ISMN) and/or cilostazol improved these outcomes. We tested whether this effect was present at 6 months, a pre-specified analysis.

**Methods:** LACI-2 assessed the feasibility, safety and proof-of-concept of one year of ISMN (40-60mg) and/or cilostazol (200 mg) using a prospective randomised open-label blinded-endpoint 2×2-factorial phase-2b design. Participants aged>30yrs had clinical lacunar stroke, compatible brain imaging and capacity to consent. Key outcomes included cognition (Diagnostic and Statistical Manual version 5-7-level, DSM5-7L), dependency, vascular events, mood, stroke impact and a global analysis of these.

**Results:** Baseline characteristics were balanced across 363 participants: median age 64 [56-72] years, females 31% and median onset-to-randomisation 79 [27-244] days. At 6 months, participants allocated to ISMN vs control had fewer vascular-cognition-dependency events (adjusted odds ratio, aOR 0.74, 95% confidence intervals 0.55-0.99) and an improved stroke impact scale score (Mann-Whitney difference, MWD - 0.15, 95% CI −0.25, −0.05) and global analysis (MWD −0.06, 95% CI −0.11, −0.01). Cilostazol vs control reduced cognitive impairment (DSM5-7-level scale, acOR 0.64, 95% CI 0.41-0.99). Combined ISMN/cilostazol vs control (n=181) had improved cognition (DSM5-7L acOR 0.40, 95% CI 0.21-0.78) and mood (Zung aMD −6.94, 95% CI −12.25, −1.64), reduced stroke impact (−0.23, 95% CI −0.37, −0.09) and a better global outcome (MWD −0.10, 95% CI −0.17, −0.03).

**Conclusions:** Improvements in cognitive and functional outcomes were seen within 6 months of ISMN and/or cilostazol.

Registration: ISRCTN14911850: https://www.isrctn.com/ISRCTN14911850

## INTRODUCTION

Arteriosclerotic cerebral small vessel disease (cSVD) contributes to 40% of dementias, 25% of ischaemic lacunar strokes, most intracerebral haemorrhage (ICH) and several conditions affecting older people including falls, apathy, fatigue, depression, delirium and incontinence. The combination of cSVD with other causes of dementia or stroke is associated with a worse outcome.^1^

cSVD is a condition involving endothelial,^2, 3^ smooth muscle, pericyte, cerebrovascular reactivity and blood brain barrier dysfunction and damage to brain networks. Hence, drugs that stabilise endothelial/blood brain barrier function might attenuate the long term clinical, cognitive and functional consequences of cSVD.^4^ Endothelial function depends on both the nitric oxide-cyclic guanosine monophosphate-phosphodiesterase-5 (NO-cAMP-PDE5) and prostacyclin-cyclic adenosine monophosphate-phosphodiesterase-3 (PGI2-cAMP-PDE3) pathways.^4^ Hence, isosorbide mononitrate (ISMN),^5^ a NO donor, and cilostazol, a PDE3-inhibitor,^6^ might improve cerebral small vessel dysfunction. Both drugs are licensed in Europe to treat large artery disease (ISMN for angina in ischaemic heart disease and cilostazol for intermittent claudication in peripheral artery disease) and so are candidates for repurposing in cSVD. Although there are no randomised data involving NO donors for stroke secondary prophylaxis, dipyridamole (a PDE3/5-inhibitor ^6^) and cilostazol each reduce stroke recurrence.^7, 8^

Our LACI-2 phase-2b 2×2 factorial trial of ISMN and cilostazol in 363 patients with prior lacunar ischaemic stroke found that both drugs were safe and could be taken for a year.^9^ After 12 months, ISMN vs no ISMN had reduced recurrent stroke, cognitive impairment, stroke impact, improved quality of life, and a global analysis of these. Cilostazol vs no cilostazol reduced dependency and mood disturbance. Combining both drugs vs neither improved most of these outcomes. We hypothesised that these effects might be evident even before one year. Here we report the early effect of ISMN and cilostazol, given either individually or together versus neither, after only six months of therapy, a planned secondary analysis based on prospectively collected data, to explore the time-frame of treatment effects and to aid in future trial design.

## METHODS

LACI-2 was an investigator-led prospective randomised open-label, blinded-endpoint (PROBE) 2×2 factorial trial conducted in 26 stroke-specialist hospitals in the United Kingdom. The trial was co-ordinated by the University of Edinburgh with randomisation and case record form programming and statistics hosted at the University of Nottingham. The protocol,^10^ statistical analysis plan (SAP),^11^ baseline data ^11, 12^ and main findings ^9^ are published. The results presented here are based on pre-specified secondary analyses using outcomes collected prospectively at 6 months and follow the main trial’s SAP.^11^ LACI-2 received approvals from the UK competent authority (Medicines and Healthcare products Regulatory Agency, MHRA), the national ethics committee (Health Research Authority, HRA 17/EM/0077) and Research and Development approvals (2017/0209/TMF) from each hospital. The trial was registered: ISRCTN14911850. We obtained written informed consent from all participants prior to enrolment and followed Good Clinical Practice Guidelines.

### Participants

We included people aged >30 years with a clinical lacunar ischaemic stroke syndrome and CT or MR brain imaging showing either a visible small subcortical/lacunar infarct or no alternative finding to account for the symptoms, i.e. no cortical infarct, haemorrhage or stroke-mimicking condition. We set no time limit between stroke and recruitment, or limitations to people with protected characteristics. Exclusions included other active brain disease, renal or hepatic impairment, dependency, and lack of capacity.^10^ Diagnostic brain imaging was collected and assessed centrally (in Edinburgh) by blinded expert review.

### Randomisation

We randomised participants to ISMN vs. no ISMN and cilostazol vs. no cilostazol, each with 1:1 ratio, using a secure internet-based web-database (https://stroke.nottingham.ac.uk/laci-2/live/laci-2_login.php). Participants with indications for, or contraindications to, one drug could be randomised to the other drug alone. We minimised on age, sex, stroke (National Institute of Health Stroke Scale, NIHSS), dependency (modified Rankin Scale, mRS), systolic BP (SBP), smoking status, time after stroke, and years of education.

### Interventions

Participants were allocated to one of four groups: i) ISMN 40-60mg daily, ii) cilostazol 200mg daily, iii) both ISMN and cilostazol, or iv) neither drug. Drugs were dispensed by hospital pharmacies and started the day after randomisation at a low dose with escalation to full or maximum tolerated dose by four weeks.^10, 13^ Participants randomised to control did not take any placebo medication. All participants continued their usual prescribed medicines including guideline-based stroke prevention, typically blood pressure and lipid lowering, clopidogrel and lifestyle advice, considered as ‘best medical therapy’.

### Clinical outcomes

The primary outcome was feasibility of recruitment and retention (defined as >95% of randomised patients retained at one year);^10^ this and adherence/tolerability to trial treatment (defined as 75% of patients taking ≥50% trial dose up to one year) are reported in the main publication.^9^ Targeted symptoms known to be caused by ISMN and cilostazol were also recorded: headache, palpitations, dizziness, loose stools, nausea, bleeding, dyspepsia, bruising and falls. The proof-of-concept clinical outcome was the composite of recurrent stroke/TIA, MI, any cognitive impairment, dependency (mRS>2) and death. These measures were recorded individually at 6 months by telephone interview (or postal questionnaire) along with modified Rankin scale (mRS, range best to worst scores, 0 to 5 ^14^), telephone Montreal cognitive assessment (tMoCA, best to worst 22 to 0), telephone interview of cognitive status-modified (TICS-M, best to worst 39 to 0), verbal fluency (animal naming, best to worst “infinity” to 0), Zung depression scale (ZDS, best to worst 0 to 100) and stroke impact scale (SIS; assesses multiple domains of participant-reported physical/cognitive function, dependency, mood and QoL).^10^ Participants who died had a score worse than any living score assigned to maintain power and prevent missing any ‘kill or cure’ effect:^15^ these scores were mRS=6, tMoCA-1, verbal fluency −1, ZDS 102.5. We calculated 7-level and 4-level ordinal cognitive outcome status (DSM5-7, DSM5-4) by operationalising the Diagnostic and Statistical Manual version 5 (DSM5)^16^ using tMoCA and TICS sub-scores.^10^ In contrast to 12 months,^9^ trail making B, EuroQoL-5D 5-level utility and EuroQoL-visual analogue scores were not collected at 6 months and so are not reported here.

### Sample size

This was based on death as the primary safety measure at 12 months.^10^ Assuming all-cause death would be 2.0%/year, the upper 95% confidence interval (CI) would be 4% in 400 patients. Hence, the trial would stop if all-cause deaths exceeded 4% on trial drugs versus no trial drugs.^10^ A sample size of 400 was considered sufficient to collect event rates for recurrent stroke, dependency and cognitive impairment for which reliable data in lacunar stroke were not available when starting.

### Statistical analysis

The analyses follow the published LACI-2 statistical analysis plan ^11, 12^ and those presented in the main publication ^9^ but with reporting here of events and status assessed at 6 months. Analyses were by intention-to-treat as per randomised allocation. We did not impute missing data (so adjusted assessments are based on complete case analyses) or adjust for multiple testing since these pre-specified secondary analyses are all hypothesis-generating.

Data are shown as number (%), median [interquartile range] or mean (standard deviation). Analyses used binary logistic regression (BLR, presented as adjusted odds ratio, aOR) for dichotomous outcomes, Cox proportional hazards regression (CPHR, adjusted hazard ratio, aHR) for time-to-event measures, ordinal logistic regression (OLR, adjusted common odds ratio, acOR) for ordered categorical outcomes or multiple linear regression (MLR, adjusted mean difference, aMD) for continuous measures. Analyses were adjusted for minimisation variables: baseline age, sex, impairment (NIHSS), dependency (mRS), systolic blood pressure (SBP), smoking status, time after stroke and years of education using raw data. Cognition outcomes were additionally adjusted for baseline Montreal cognitive assessment (MoCA). 95% CI are given. Fisher’s exact test (FET) was used in *post hoc* unadjusted binary analyses to compare events with limited numbers where BLR could not be calculated.

The Wei-Lachin test was used to provide unadjusted global analyses of:^17^ i) a global clinical outcome based on recurrent ordinal stroke, ordinal MI, DSM5-7L, ordinal mRS, EQ-5D-5L utility, ZDS and death; and ii) a global stroke impact scale (SIS) score, using individual SIS domain scores. Both are reported as Mann-Whitney difference (MWD).^17^ Ordinal stroke was defined as the occurrence of stroke modified using the mRS to determine the severity of the event.^15^

## RESULTS

### Study participants

Recruitment ran between 5th February 2018 and 31st May 2021 with a short suspension during March-July 2020 due to COVID-19 lockdown. Of the planned 400 participants, 363 were recruited (see CONSORT diagram in main publication ^9^), the deficit reflecting the lost recruitment due to COVID-19. Patient characteristics were well balanced at baseline and typical of lacunar stroke (Table 1):^9, 11, 12^ median age 64 [57-72] years; more males (246, 69%); stroke onset to randomisation 79 [27-247] days; median NIHSS 0.0 [0.0-2.0] and hypertension (252, 71%). By 6 months, 5 participants had withdrawn (ISMN 1, cilostazol 2, both 2), 7 were lost to follow-up (ISMN 5, cilostazol 1, both 1), 9 had refused treatment (cilostazol 5, both 3, control 1) and 2 had died (ISMN 1, control 1).^9^

**TABLE 1.**
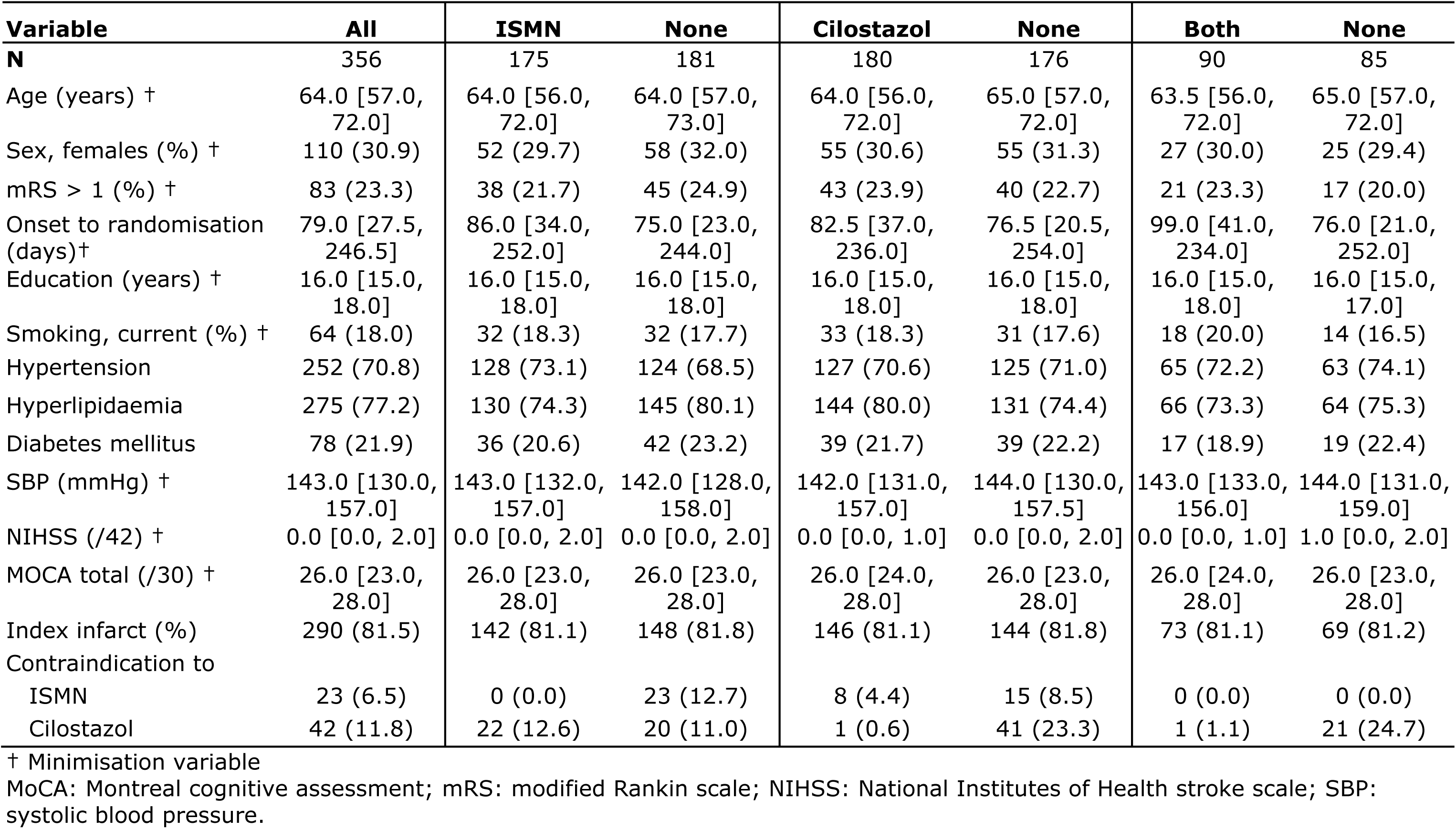
Baseline characteristics in participants with outcome data at 6 months. Data are number (%) or median [interquartile range].

| **Variable** | **All** | **ISMN** | **None** | **Cilostazol** | **None** | **Both** | **None** |
| --- | --- | --- | --- | --- | --- | --- | --- |
| **N** | 356 | 175 | 181 | 180 | 176 | 90 | 85 |
| Age (years) † | 64.0 [57.0, 72.0] | 64.0 [56.0, 72.0] | 64.0 [57.0, 73.0] | 64.0 [56.0, 72.0] | 65.0 [57.0, 72.0] | 63.5 [56.0, 72.0] | 65.0 [57.0, 72.0] |
| Sex, females (%) † | 110 (30.9) | 52 (29.7) | 58 (32.0) | 55 (30.6) | 55 (31.3) | 27 (30.0) | 25 (29.4) |
| mRS > 1 (%) † | 83 (23.3) | 38 (21.7) | 45 (24.9) | 43 (23.9) | 40 (22.7) | 21 (23.3) | 17 (20.0) |
| Onset to randomisation (days)† | 79.0 [27.5, 246.5] | 86.0 [34.0, 252.0] | 75.0 [23.0, 244.0] | 82.5 [37.0, 236.0] | 76.5 [20.5, 254.0] | 99.0 [41.0, 234.0] | 76.0 [21.0, 252.0] |
| Education (years) † | 16.0 [15.0, 18.0] | 16.0 [15.0, 18.0] | 16.0 [15.0, 18.0] | 16.0 [15.0, 18.0] | 16.0 [15.0, 18.0] | 16.0 [15.0, 18.0] | 16.0 [15.0, 17.0] |
| Smoking, current (%) † | 64 (18.0) | 32 (18.3) | 32 (17.7) | 33 (18.3) | 31 (17.6) | 18 (20.0) | 14 (16.5) |
| Hypertension | 252 (70.8) | 128 (73.1) | 124 (68.5) | 127 (70.6) | 125 (71.0) | 65 (72.2) | 63 (74.1) |
| Hyperlipidaemia | 275 (77.2) | 130 (74.3) | 145 (80.1) | 144 (80.0) | 131 (74.4) | 66 (73.3) | 64 (75.3) |
| Diabetes mellitus | 78 (21.9) | 36 (20.6) | 42 (23.2) | 39 (21.7) | 39 (22.2) | 17 (18.9) | 19 (22.4) |
| SBP (mmHg) † | 143.0 [130.0, 157.0] | 143.0 [132.0, 157.0] | 142.0 [128.0, 158.0] | 142.0 [131.0, 157.0] | 144.0 [130.0, 157.5] | 143.0 [133.0, 156.0] | 144.0 [131.0, 159.0] |
| NIHSS (/42) † | 0.0 [0.0, 2.0] | 0.0 [0.0, 2.0] | 0.0 [0.0, 2.0] | 0.0 [0.0, 1.0] | 0.0 [0.0, 2.0] | 0.0 [0.0, 1.0] | 1.0 [0.0, 2.0] |
| MOCA total (/30) † | 26.0 [23.0, 28.0] | 26.0 [23.0, 28.0] | 26.0 [23.0, 28.0] | 26.0 [24.0, 28.0] | 26.0 [23.0, 28.0] | 26.0 [24.0, 28.0] | 26.0 [23.0, 28.0] |
| Index infarct (%) | 290 (81.5) | 142 (81.1) | 148 (81.8) | 146 (81.1) | 144 (81.8) | 73 (81.1) | 69 (81.2) |
| Contraindication to |  |  |  |  |  |  |  |
| ISMN | 23 (6.5) | 0 (0.0) | 23 (12.7) | 8 (4.4) | 15 (8.5) | 0 (0.0) | 0 (0.0) |
| Cilostazol | 42 (11.8) | 22 (12.6) | 20 (11.0) | 1 (0.6) | 41 (23.3) | 1 (1.1) | 21 (24.7) |

† Minimisation variable
MoCA: Montreal cognitive assessment; mRS: modified Rankin scale; NIHSS: National Institutes of Health stroke scale; SBP: systolic blood pressure.

### Adherence

At six months, 87.6% of participants were taking assigned treatment, a figure that was similar across the treatment groups (eTable 1 in ^9^). 68.1% of participants were taking 50% or more of ISMN and/or cilostazol doses.

### Clinical outcomes at 6 months

By 6 months, few stroke recurrences (8) and deaths (2) and no MIs had occurred (Table 2) so the proof-of-concept primary clinical outcome (composite of binary events) was driven primarily by cognitive impairment (DSM5-7L>0) and dependency (mRS>2) events. The primary clinical outcome was reduced with ISMN vs no ISMN (aOR 0.74, 95% CI 0.55-0.99, p=0.040; Table 2, Figure 1a) but not cilostazol (Figure 1b). A tendency to fewer events was present with combined ISMN and cilostazol vs neither (aOR 0.71, 95% CI 0.45-1.13, p=0.15; Figure 1c). ISMN was associated with reduced stroke/TIA recurrence (Table 2, p=0.002).

**FIGURE 1.**
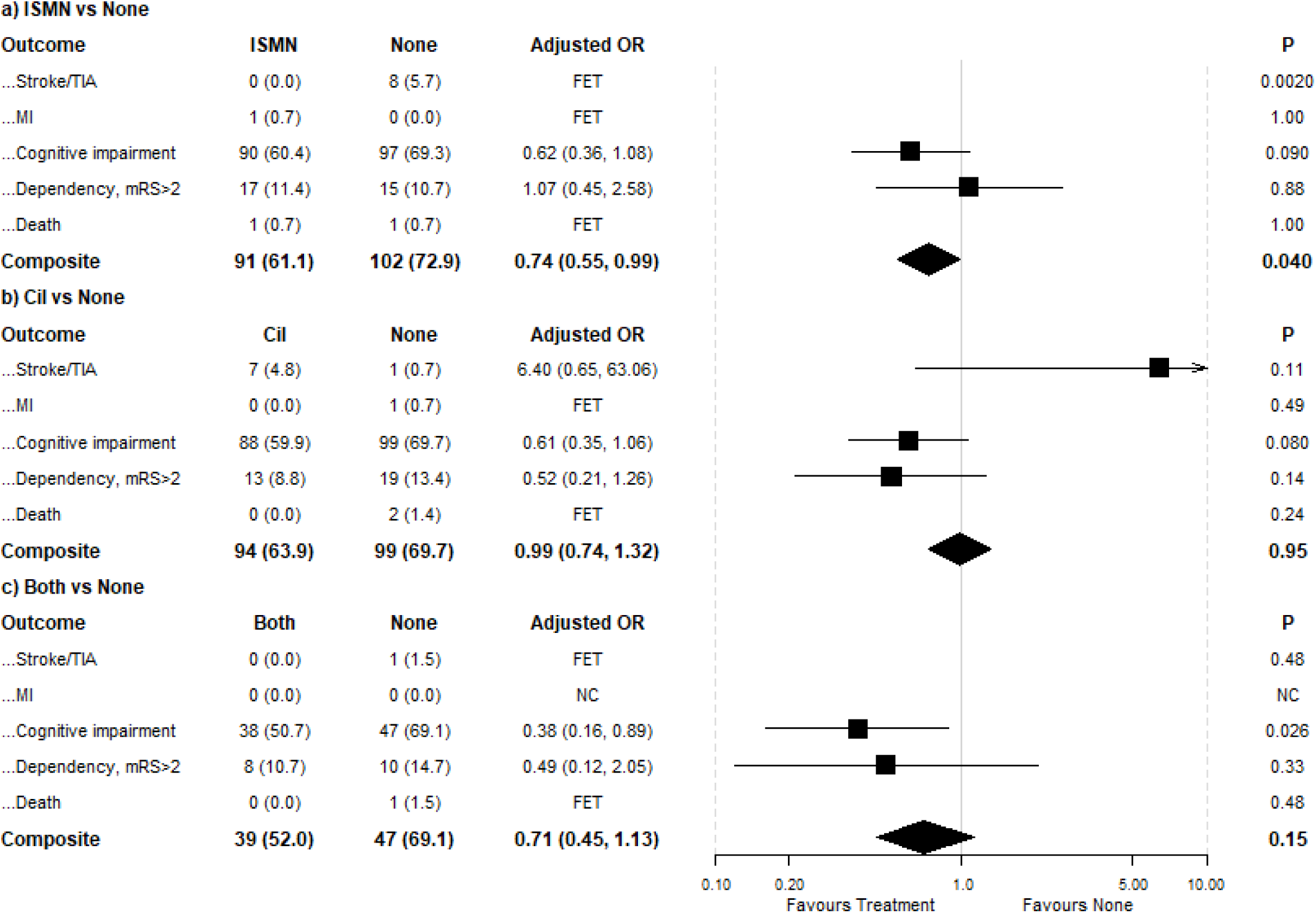
Effect of interventions on composite outcome. Forest plots of composite outcome (recurrent stroke or TIA, MI, any cognitive impairment, dependency mRS >2, death) by randomised treatment and subgroups: Data are adjusted odds ratio (aOR, 95% confidence intervals, 95% CI) with analysis by adjusted binary logistic regression. Adjustment for baseline characteristics including age, sex, onset time to randomisation, systolic blood pressure, pre-stroke mRS, MoCA and education level except for the Fisher’s exact test (FET) analysis. Stroke and MI events were too infrequent to calculate aOR.

**TABLE 2.** Outcome measures at 6 months. Data are number (%) or median [interquartile range]. Comparisons by binary logistic regression, Fisher’s exact test, ordinal logistic regression, or multiple linear regression.

| Outcomes | N | All | Any ISMN | None | aOR/acOR/aHR/aMD (95% CI) | p | Any Cil | None | aOR/acOR/aHR/aMD (95% CI) | p | Both | None | aOR/acOR/aHR/aMD (95% CI) | p |
| --- | --- | --- | --- | --- | --- | --- | --- | --- | --- | --- | --- | --- | --- | --- |
| **Number of patients** |  | **356** | **175** | **181** |  |  | **180** | **176** |  |  | **90** | **91** |  |  |
| Composite | 289 | 193 (66.8) | 91 (61.1) | 102 (72.9) | 0.74 (0.55, 0.99) | 0.040 | 94 (63.9) | 99 (69.7) | 0.99 (0.74, 1.32) | 0.95 | 39 (52.0) | 47 (69.1) | 0.71 (0.45, 1.13) | 0.15 |
| Stroke/TIA | 289 | 8 (2.8) | 0 (0.0) | 8 (5.7) | / | 0.002 | 7 (4.8) | 1 (0.7) | 6.40 (0.65, 63.06) | 0.11 | 0 (0.0) | 1 (1.5) | / | 0.48 |
| MI | 289 | 1 (0.3) | 1 (0.7) | 0 (0.0) | / | 1.00 | 0 (0.0) | 1 (0.7) | / | 0.49 | 0 (0.0) | 0 (0.0) | - | - |
| Cognitive impairment | 289 | 187 (64.7) | 90 (60.4) | 97 (69.3) | 0.62 (0.36, 1.08) | 0.090 | 88 (59.9) | 99 (69.7) | 0.61 (0.35, 1.06) | 0.080 | 38 (50.7) | 47 (69.1) | 0.38 (0.16, 0.89) | 0.026 |
| Dependency, mRS>2 | 289 | 32 (11.1) | 17 (11.4) | 15 (10.7) | 1.07 (0.45, 2.58) | 0.88 | 13 (8.8) | 19 (13.4) | 0.52 (0.21, 1.26) | 0.14 | 8 (10.7) | 10 (14.7) | 0.49 (0.12, 2.05) | 0.33 |
| Death | 289 | 2 (0.7) | 1 (0.7) | 1 (0.7) | - | - | 0 (0.0) | 2 (1.4) | - | - | 0 (0.0) | 1 (1.5) | / | / |
| Sensitivity: Composite (any) | 312 | 216 (69.2) | 98 (62.8) | 118 (75.6) | 0.74 (0.56, 0.98) | 0.035 | 102 (65.8) | 114 (72.6) | 0.91 (0.69, 1.20) | 0.52 | 43 (54.4) | 59 (73.8) | 0.65 (0.42, 0.99) | 0.044 |
| Sensitivity: Stroke or TIA (any) | 356 | 14 (3.9) | 0 (0.0) | 14 (7.7) | - | - | 9 (5.0) | 5 (2.8) | 1.68 (0.51, 5.57) | 0.39 | 0 (0.0) | 5 (5.5) | - | - |
| Sensitivity: MI (any) | 356 | 1 (0.3) | 1 (0.6) | 0 (0.0) | - | - | 0 (0.0) | 1 (0.6) | - | - | 0 (0.0) | 0 (0.0) | - | - |
| Cognition |  |  |  |  |  |  |  |  |  |  |  |  |  |  |
| Cognition, 7-level | 356 | - | - | - | 0.66 (0.43, 1.02) | 0.059 | - | - | 0.64 (0.41, 0.99) | 0.043 | - | - | 0.40 (0.21, 0.78) | 0.007 |
| Normal | 306 | 105 (34.3) | 61 (38.6) | 44 (29.7) |  |  | 60 (39.0) | 45 (29.6) |  |  | 38 (47.5) | 22 (29.7) |  |  |
| Minor, single domain | 306 | 84 (27.5) | 41 (25.9) | 43 (29.1) |  |  | 39 (25.3) | 45 (29.6) |  |  | 16 (20.0) | 20 (27.0) |  |  |
| Minor, multi-domain | 306 | 45 (14.7) | 22 (13.9) | 23 (15.5) |  |  | 26 (16.9) | 19 (12.5) |  |  | 12 (15.0) | 9 (12.2) |  |  |
| Major, Mild | 306 | 62 (20.3) | 29 (18.4) | 33 (22.3) |  |  | 27 (17.5) | 35 (23.0) |  |  | 13 (16.3) | 19 (25.7) |  |  |
| Major, Moderate | 306 | 8 (2.6) | 4 (2.5) | 4 (2.7) |  |  | 2 (1.3) | 6 (3.9) |  |  | 1 (1.3) | 3 (4.1) |  |  |
| Major, Severe | 306 | 0 (0.0) | 0 (0.0) | 0 (0.0) |  |  | 0 (0.0) | 0 (0.0) |  |  | 0 (0.0) | 0 (0.0) |  |  |
| Died | 306 | 2 (0.7) | 1 (0.6) | 1 (0.7) |  |  | 0 (0.0) | 2 (1.3) |  |  | 0 (0.0) | 1 (1.4) |  |  |
| Cognition, 4-level | 356 | - | - | - | 0.66 (0.42, 1.03) | 0.066 | - | - | 0.59 (0.37, 0.92) | 0.020 | - | - | 0.39 (0.20, 0.76) | 0.006 |
| Normal | 306 | 105<br>(34.3) | 61<br>(38.6) | 44<br>(29.7) |  |  | 60<br>(39.0) | 45<br>(29.6) |  |  | 38<br>(47.5) | 22<br>(29.7) |  |  |
| Minor | 306 | 129<br>(42.2) | 63<br>(39.9) | 66<br>(44.6) |  |  | 65<br>(42.2) | 64<br>(42.1) |  |  | 28<br>(35.0) | 29<br>(39.2) |  |  |
| Dementia | 306 | 70<br>(22.9) | 33<br>(20.9) | 37<br>(25.0) |  |  | 29<br>(18.8) | 41<br>(27.0) |  |  | 14<br>(17.5) | 22<br>(29.7) |  |  |
| Death | 306 | 2 (0.7) | 1 (0.6) | 1 (0.7) |  |  | 0 (0.0) | 2 (1.3) |  |  | 0 (0.0) | 1 (1.4) |  |  |
| Memory/thinking problem apparent to participant | 315 | 209<br>(66.3) | 95<br>(60.9) | 114<br>(71.7) | 0.61 (0.37, 1.02) | 0.059 | 105<br>(66.5) | 104<br>(66.2) | 0.99 (0.60, 1.64) | 0.97 | 49<br>(61.3) | 58<br>(71.6) | 0.55 (0.26, 1.16) | 0.12 |
| tMoCA (/22) | 311 | 18.2<br>(3.3) | 18.3<br>(3.3) | 18.1<br>(3.2) | 0.14 (-0.51, 0.79) | 0.68 | 18.4<br>(3.0) | 17.9<br>(3.6) | 0.46 (-0.19, 1.12) | 0.16 | 18.7<br>(3.1) | 18.1<br>(3.6) | 0.60 (-0.34, 1.54) | 0.21 |
| TICS-M (/39) | 315 | 25.9<br>(4.9) | 26.1<br>(4.9) | 25.8<br>(4.8) | 0.10 (-0.88, 1.09) | 0.84 | 26.3<br>(4.4) | 25.5<br>(5.2) | 0.82 (-0.17, 1.80) | 0.10 | 26.6<br>(4.7) | 25.5<br>(5.4) | 0.96 (-0.47, 2.39) | 0.19 |
| Verbal fluency (animal naming) | 328 | 19.6<br>(6.9) | 20.2<br>(7.5) | 19.0<br>(6.2) | 1.07 (-0.30, 2.43) | 0.13 | 19.8<br>(6.6) | 19.4<br>(7.2) | 0.22 (-1.14, 1.58) | 0.76 | 20.3<br>(7.1) | 18.7<br>(6.4) | 1.36 (-0.38, 3.10) | 0.13 |
| Dementia, clinical diagnosis (SAE) | 356 | 0 (0.0) | 0 (0.0) | 0 (0.0) | - | - | 0 (0.0) | 0 (0.0) | - | - | 0 (0.0) | 0 (0.0) | - | - |
| Clinical |  |  |  |  |  |  |  |  |  |  |  |  |  |  |
| Extracranial bleeding | 356 | 2 (0.6) | 2 (1.1) | 0 (0.0) | / | 0.24 | 1 (0.6) | 1 (0.6) | / | 1.00 | 1 (1.1) | 0 (0.0) | / | 0.49 |
| mRS | 320 | 1.0 [1.0, 2.0] | 1.0 [1.0, 2.0] | 1.0 [1.0, 2.0] | 0.81 (0.53, 1.22) | 0.30 | 1.0 [1.0, 2.0] | 1.0 [1.0, 2.0] | 0.77 (0.51, 1.17) | 0.22 | 1.0 [1.0, 2.0] | 2.0 [1.0, 2.0] | 0.61 (0.33, 1.10) | 0.099 |
| Disposition | 320 | 1.0 [1.0, 1.0] | 1.0 [1.0, 1.0] | 1.0 [1.0, 1.0] | 0.60 (0.05, 7.64) | 0.70 | 1.0 [1.0, 1.0] | 1.0 [1.0, 1.0] | 0.41 (0.04, 4.87) | 0.48 | 1.0 [1.0, 1.0] | 1.0 [1.0, 1.0] | / | / |
| Zung depression scale (/102.5) | 316 | 50.5<br>(17.5) | 48.6<br>(17.0) | 52.3<br>(17.9) | -3.16 (-6.72, 0.40) | 0.082 | 49.2<br>(17.0) | 51.7<br>(18.0) | -2.96 (-6.54, 0.61) | 0.10 | 48.1<br>(18.0) | 54.2<br>(19.4) | -6.94 (-12.25, -1.64) | 0.010 |
| Global | 306 | - | - | - | -0.06 (-0.11, -0.01) | 0.025 | - | - | -0.05 (-0.10, 0.00) | 0.076 | - | - | -0.10 (-0.17, -0.03) | 0.008 |
| SIS |  |  |  |  |  |  |  |  |  |  |  |  |  |  |
| Strength (/100) | 307 | 77.6<br>(22.6) | 80.1<br>(21.8) | 75.3<br>(23.2) | 3.87 (-0.64, 8.37) | 0.093 | 78.4<br>(21.5) | 77.0<br>(23.7) | 1.08 (-3.44, 5.61) | 0.64 | 79.4<br>(20.4) | 73.4<br>(23.6) | 4.44 (-1.65, 10.53) | 0.15 |
| Memory (/100) | 317 | 83.8<br>(20.2) | 87.0<br>(17.5) | 80.6<br>(22.2) | 5.86 (1.77, 9.95) | 0.005 | 85.9<br>(18.0) | 81.6<br>(22.0) | 4.55 (0.45, 8.66) | 0.030 | 88.9<br>(14.3) | 78.4<br>(23.4) | 11.66 (6.22, 17.09) | <0.001 |
| Emotion (/100) | 316 | 72.1<br>(20.3) | 74.5<br>(20.0) | 69.8<br>(20.3) | 3.95 (-0.03, 7.92) | 0.052 | 75.3<br>(19.7) | 69.0<br>(20.4) | 6.97 (2.98, 10.97) | <0.001 | 75.7<br>(20.8) | 65.1<br>(20.9) | 11.74 (5.98, 17.50) | <0.001 |
| Communication (/100) | 316 | 90.3<br>(16.7) | 92.8<br>(13.8) | 87.8<br>(18.9) | 4.66 (1.23, 8.10) | 0.0077 | 92.2<br>(14.6) | 88.4<br>(18.4) | 3.82 (0.38, 7.27) | 0.030 | 93.9<br>(11.1) | 85.4<br>(20.0) | 9.05 (4.35, 13.76) | <0.001 |
| Activities of daily living (/100) | 314 | 87.5 (18.6) | 89.4 (16.9) | 85.7 (20.0) | 3.05 (-0.66, 6.75) | 0.11 | 89.3 (16.1) | 85.7 (20.7) | 3.36 (-0.36, 7.08) | 0.077 | 90.5 (14.8) | 83.3 (22.0) | 7.19 (1.86, 12.52) | 0.009 |
| Mobility (/100) | 315 | 83.7 (20.8) | 85.9 (19.4) | 81.5 (21.9) | 3.51 (-0.48, 7.50) | 0.085 | 85.4 (18.8) | 82.0 (22.5) | 3.34 (-0.67, 7.34) | 0.10 | 86.3 (18.1) | 78.7 (23.7) | 7.25 (1.51, 12.98) | 0.013 |
| Hand (/100) | 311 | 81.1 (26.0) | 83.1 (24.6) | 79.1 (27.3) | 2.89 (-2.30, 8.08) | 0.28 | 83.8 (23.0) | 78.5 (28.4) | 4.98 (-0.24, 10.20) | 0.061 | 85.8 (21.0) | 76.6 (29.3) | 8.63 (1.33, 15.92) | 0.020 |
| Role (/100) | 289 | 79.8 (25.3) | 82.8 (24.6) | 76.8 (25.7) | 5.16 (0.04, 10.28) | 0.048 | 82.1 (23.0) | 77.4 (27.3) | 4.56 (-0.58, 9.70) | 0.082 | 84.2 (23.3) | 73.5 (28.3) | 10.71 (3.58, 17.84) | 0.004 |
| Recovery (/100) | 315 | 77.5 (22.6) | 81.2 (20.3) | 74.0 (24.2) | 6.21 (1.84, 10.57) | 0.005 | 80.1 (21.0) | 75.0 (23.9) | 4.77 (0.39, 9.15) | 0.033 | 82.3 (19.3) | 70.2 (25.2) | 11.41 (5.41, 17.41) | <0.001 |
| Global SIS | 289 | - | - | - | -0.15 (-0.25, -0.05) | 0.004 | - | - | -0.09 (-0.19, 0.01) | 0.084 | - | - | -0.23 (-0.37, -0.09) | 0.002 |

/: Fisher's exact test so no point estimate (95% CI).
aHR: adjusted hazard ratio; aMD: adjusted mean difference; acOR: adjusted common odds ratio; aOR: adjusted odds ratio; mRS: modified Rankin scale; SIS: stroke impact scale; TICS-M: telephone interview of cognitive status-modified; tMoCA: telephone Montreal cognitive assessment.
Sensitivity analyses based on presence of any data within a composite outcome.

No cases of dementia had occurred by 6 months. The 7-level cognitive outcome (DSM5-7) was non-significantly reduced with ISMN (acOR 0.66, 95% CI 0.43-1.02, p=0.059; Table 2, Figure 2a) and significantly with cilostazol (acOR 0·64, 95% CI 0·41-0·99, p=0.043; Figure 2b) and combined ISMN and cilostazol (acOR 0.40, 95% CI 0.21-0.78, p=0.007; Figure 2c). Similar results were seen for the DSM5-4 level (Table 2). Neither intervention, given separately or together, altered individual measures of cognition including tMoCA, TICS-M or verbal fluency. Similarly, mRS and disposition did not differ between the treatment groups. The combination of ISMN and cilostazol was associated with a significantly lower Zung depression scale score, aMD - 6.94 (95% CI −12.25 to –1.64, p=0.01; Table 2). Global analysis of recurrent ordinal stroke, ordinal MI, DSM5-7, ordinal mRS, EQ-5D-5L utility, ZDS and death favoured ISMN (MWD −0.06, 95% CI −0.11 to −0.11, p=0.025), cilostazol (MWD −0.05, 95% CI −0.10 to 0.00, p=0.076) and their combination (MWD −0.10, 95% CI −0.17 to 0.03, p=0.008).

**FIGURE 2.**
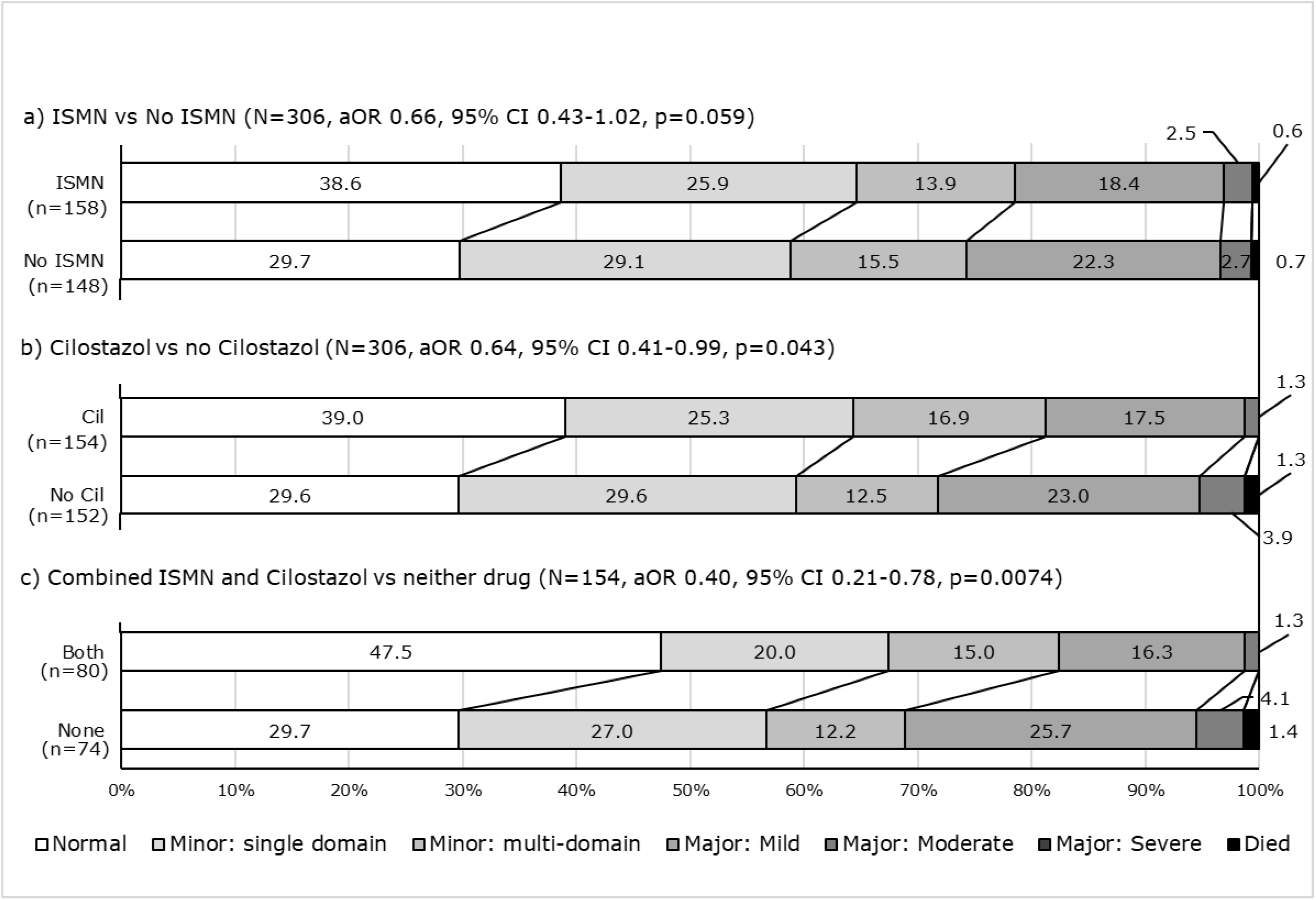
Effect of interventions on DSM5 7-level cognition scale. Data are adjusted common odds ratio (acOR, 95% confidence intervals, 95% CI) with analysis by adjusted ordinal logistic regression. Adjustment for baseline characteristics including age, sex, onset time to randomisation, systolic blood pressure, pre-stroke mRS, MoCA and education level.

ISMN was associated with higher (better) SIS scores in all domains ranging between 2.89 (hand - non-significant) to 6.21 (recovery - significant) points, and so a better global analysis of SIS (MWD −0.15, 95% CI −0.25 to −0.05, p=0.004; Table 2). Similarly, cilostazol was associated with better SIS scores although the global analysis was non-significant (MWD −0.09, 95% CI −0.19, 0.01, p=0.084). The combination of ISMN and cilostazol was associated with higher SIS scores in all domains (except strength) and a superior global analysis (MWD −0.23, 95% CI −0.37 to −0.09, p=0.002; Table 2).

Subgroup analyses by sex were presented in the main publication;^9^ none showed a treatment-sex interaction and so these are not repeated here for the 6-month data. Information on race-ethnicity were not collected.

### Safety by 6 months

Serious adverse events did not differ during treatment between ISMN vs no ISMN (12 [6.6%] vs 11 [6.0%], FET 2p=0.83), cilostazol vs no cilostazol (26 [14.3%] vs 17 [9.4%], FET 2p=0.19) or combined drugs vs no drugs (13 [14.3%] vs 10 [11.0%], FET 2p=0.66). For targeted symptoms occurring by 6 months of therapy, ISMN was associated with increased headache (121 [69%] vs 87 [48%], 2p<0.001) and potentially less bleeding (16 [9%] vs 27 [15%], 2p=0.080). Cilostazol was associated with increased palpitations (46 [25%] vs 31 [17%], 2p=0.059) and loose stools (102 [56%] vs 53 [29%], 2p<0.001). Combination therapy was associated with increased headache (62 [68%] vs 41 [45%], 2p=0.002) and loose stools (46 [51%] vs 28 [31%], 2p=0.007). None of these symptoms led to limitations on normal activities of living.

### Comparison of findings at 6 versus 12 months

Table 3 shows a qualitative comparison of the findings at 6 and 12 months. All analyses, whether significant, non-significant or neutral, had point estimates in the appropriate direction for benefit. In general, findings at 6 and 12 months were qualitatively similar with two caveats: first, that some between-treatment differences for outcomes at 6 months were smaller than at 12 months and, second, the limited number of binary events at 6 months meant that analyses were underpowered.

**TABLE 3.**
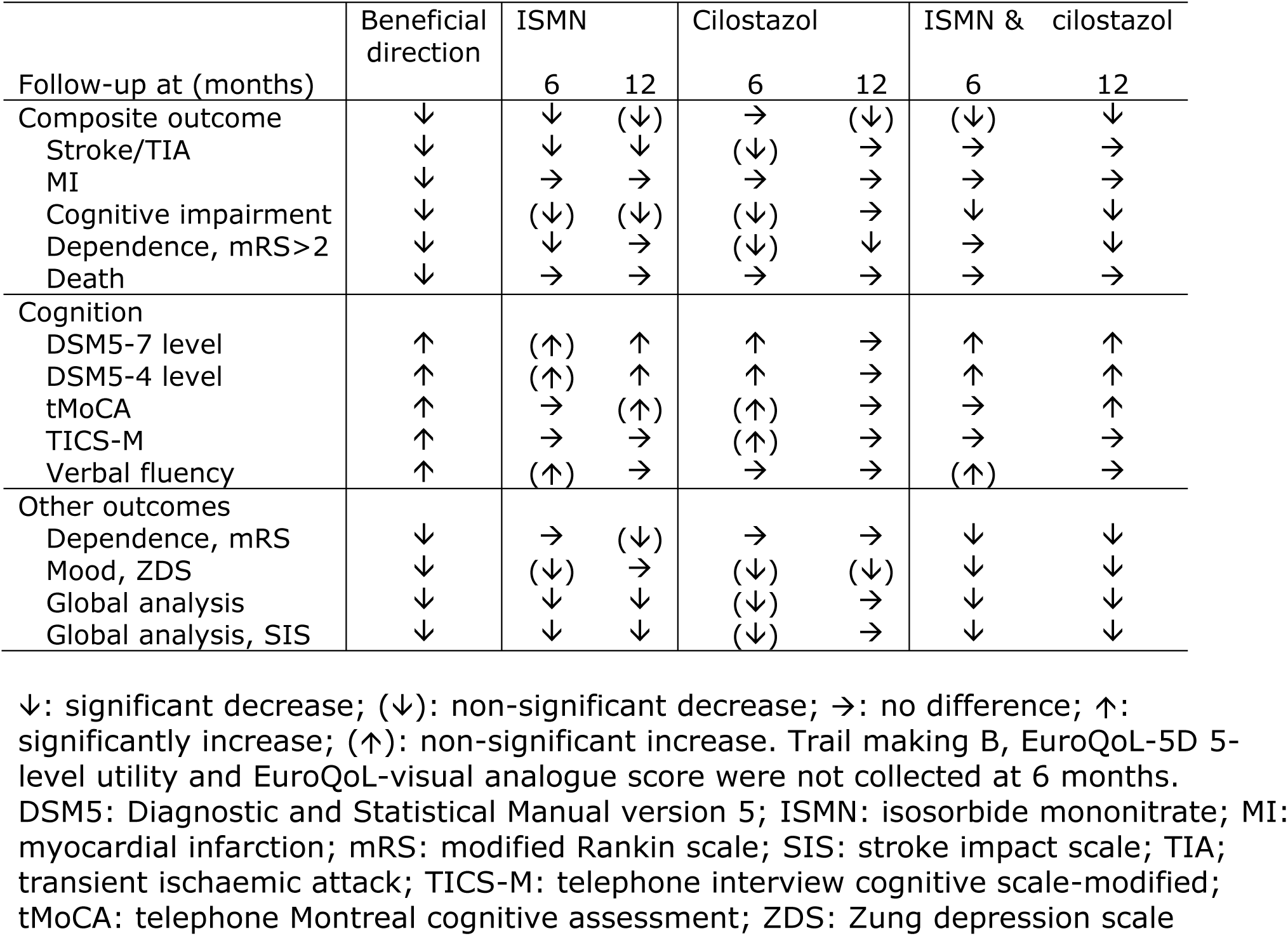
Qualitative comparison of findings at 6 versus 12 months.

| Follow-up at (months) | Beneficial direction | ISMN |  | Cilostazol |  | ISMN & cilostazol |  |
| --- | --- | --- | --- | --- | --- | --- | --- |
|  |  | 6 | 12 | 6 | 12 | 6 | 12 |
| Composite outcome | ↓ | ↓ | (↓) | → | (↓) | (↓) | ↓ |
| Stroke/TIA | ↓ | ↓ | ↓ | (↓) | → | → | → |
| MI | ↓ | → | → | → | → | → | → |
| Cognitive impairment | ↓ | (↓) | (↓) | (↓) | → | ↓ | ↓ |
| Dependence, mRS>2 | ↓ | ↓ | → | (↓) | ↓ | → | ↓ |
| Death | ↓ | → | → | → | → | → | → |
| Cognition |  |  |  |  |  |  |  |
| DSM5-7 level | ↑ | (↑) | ↑ | ↑ | → | ↑ | ↑ |
| DSM5-4 level | ↑ | (↑) | ↑ | ↑ | → | ↑ | ↑ |
| tMoCA | ↑ | → | (↑) | (↑) | → | → | ↑ |
| TICS-M | ↑ | → | → | (↑) | → | → | → |
| Verbal fluency | ↑ | (↑) | → | → | → | (↑) | → |
| Other outcomes |  |  |  |  |  |  |  |
| Dependence, mRS | ↓ | → | (↓) | → | → | ↓ | ↓ |
| Mood, ZDS | ↓ | (↓) | → | (↓) | (↓) | ↓ | ↓ |
| Global analysis | ↓ | ↓ | ↓ | (↓) | → | ↓ | ↓ |
| Global analysis, SIS | ↓ | ↓ | ↓ | (↓) | → | ↓ | ↓ |

↓: significant decrease; (↓): non-significant decrease; →: no difference; ↑: significantly increase; (↑): non-significant increase. Trail making B, EuroQoL-5D 5-level utility and EuroQoL-visual analogue score were not collected at 6 months. DSM5: Diagnostic and Statistical Manual version 5; ISMN: isosorbide mononitrate; MI: myocardial infarction; mRS: modified Rankin scale; SIS: stroke impact scale; TIA; transient ischaemic attack; TICS-M: telephone interview cognitive scale-modified; tMoCA: telephone Montreal cognitive assessment; ZDS: Zung depression scale

## DISCUSSION

This pre-planned secondary analysis of the LACI-2 trial shows that many of the positive findings seen after 12 months of ISMN and/or cilostazol ^9^ were either present at 6 months or were developing by this time. This included an extended composite vascular outcome, the DSM5 7-level ordinal cognition scale, stroke impact scale components and global analyses of all these.

We originally identified ISMN and cilostazol as potential agents that might modify cSVD development and its clinical consequences following an extensive review in 2015.^4^ cSVD involves endothelial dysfunction including blood brain barrier dysfunction and agents that enhance the NO-cAMP-PDE5 and PGI2-cAMP-PDE3 pathways might be expected to stabilise endothelium, blood brain barrier and reduce secondary effects on brain networks and so attenuate the clinical manifestations of cSVD.^4, 18^ The vasodilatory, antiplatelet and antileukocyte effects of ISMN and cilostazol develop rapidly so it not surprising that effects on clinical measures become apparent within six months. Positive vascular dementia trials of a similar size have been published previously on cognition at 6 months, e.g. with donepezil.^19, 20^

The strengths of this 6-month analysis is that it was preplanned, the 6-month data were collected prospectively and the analyses followed the plan used in the main publication. Further, the findings were consistent across multiple outcomes with point estimates all in favour of efficacy with ISMN and, especially, with combined ISMN and cilostazol (Table 3).

Weaknesses are, first, that the trial was open-label and so, in principle, findings may have been biased by knowledge of assignment. Nevertheless, telephone follow-ups were performed blinded to treatment assignment and participants/carers were asked not to say their treatment if remembered. Second, although ISMN might improve outcomes, the results were less in favour of cilostazol despite multiple trials in East Asia suggesting it reduces stroke recurrence.^8^ It is possible that longer treatment than either 6 or 12 months is required to see the full effects of cilostazol since benefits were only seen in trials with several years of treatment. Third, the trial was disrupted by COVID-19 and it is likely that the intended sample size of 400 would have been reached if lockdowns had not prevented recruitment. As such, the actual sample size of 363 means analyses achieved less power than intended. Fourth, there were a limited number of events for stroke and death and no MIs (as expected in lacunar stroke), and the primary proof of concept analysis involving composite events was primarily driven by data on cognitive impairment and dependency. Last, the trial was primarily designed to assess feasibility, tolerability, safety and proof of concept so was not formally powered to assess efficacy or subgroup or drug interactions.

In summary ISMN and/or cilostazol, especially when taken together, reduced composite events, cognitive impairment, mood disorders and stroke impact at 6 months in a broadly similar pattern to the main findings at 12 months.^9^ The LACI-2 findings need to be confirmed and the phase-3 LACI-3 trial (ISRCTN44436843) is underway assessing efficacy (as cognitive impairment, DSM5-7 ^16^) over 18 months of treatment. A sister phase-2 trial to LACI-2, CVD-Cog (IRAS 1011543), is assessing the effect of the same agents given for 6 months in patients with non-lacunar ischaemic stroke who also have radiological evidence of cSVD. The LACI-2 six-month findings suggest that 6-months of treatment and follow-up may be sufficient to demonstrate proof-of-concept in phase-2b/c trials.

## Data Availability

The CI, with approval from the TSC as necessary, will consider all reasonable requests to share individual participant data on provision of a protocol detailing aims, hypotheses, analyses, tables, figures and publication plan. Where possible, we will perform the analyses; alternatively, de-identified data and a data dictionary will be provided for remote analyses, subject to signed data access agreement.

## ACKNOWLEDGEMENTS

We thank the patients and their families for participating in LACI-2, all sites and investigators that made LACI-2 possible (Appendix). PMB and JMW had full access to all of the data in the study and jointly take responsibility for the integrity of the data and the accuracy of the data analysis. PMB and JMW contributed equally to this work.

## DATA SHARING

See supplement.

## Non-standard abbreviations and acronyms

acOR: adjusted common odds ratio
aOR: adjusted odds ratio
cSVD: cerebral small vessel disease
DSM5-7L: Diagnostic and Statistical Manual version 5-7-level
ISMN: isosorbide-mononitrate
LACI-2: LACunar Intervention Trial-2
MWD: Mann-Whitney difference
NO-cAMP-PDE5: nitric oxide-cyclic guanosine monophosphate-phosphodiesterase-5
PGI2-cAMP-PDE3: prostacyclin-cyclic adenosine monophosphate-phosphodiesterase-3
PROBE: prospective randomised open-label, blinded-endpoint

